# Accuracy and Consistency of Online Chat-based Artificial Intelligence Platforms in Answering Patients’ Questions About Heart Failure

**DOI:** 10.1101/2023.09.12.23295452

**Authors:** Elie Kozaily, Mabelissa Geagea, Ecem Raziye Akdogan, Jessica Atkins, Mohamed B. Elshazly, Maya Guglin, Ryan J Tedford, Ramsey M. Wehbe

**Author notes:** **Address for Correspondence: Ramsey M Wehbe, MD, MSAI** Assistant Professor of Medicine, Division of Cardiology &, Biomedical Informatics Center (BMIC), Lead AI Clinical Scientist, MUSC AI Hub, Medical University of South Carolina, 22 WestEdge Street, Suite 200, Room WG213E, Charleston, SC 29403, Twitter: @ramseywehbemd.

## Abstract

**Background:** Heart failure (HF) is a prevalent condition associated with significant morbidity. Patients may have questions that they feel embarrassed to ask or will face delays awaiting responses from their healthcare providers which may impact their health behavior. We aimed to investigate the potential of chat-based artificial intelligence (AI) platforms in complementing the delivery of patient-centered care.

**Methods:** Using online patient forums and physician experience, we created 30 questions related to diagnosis, management and prognosis of HF. The questions were posed to two artificial intelligence (AI) chatbots (OpenAI’s ChatGPT-3.5 and Google’s Bard). Each set of answers was evaluated by two HF experts, independently and blinded to each other, for accuracy (adequacy of content) and consistency of content.

**Results:** ChatGPT provided mostly appropriate answers (27/30, 90%) and showed a high degree of consistency (93%). Bard provided a similar content in its answers and thus was evaluated only for adequacy (23/30, 77%). The two HF experts’ grades were concordant in 83% and 67% of the questions for ChatGPT and Bard, respectively. Both platforms suffered from issues related to “hallucination” of facts and/or difficulty with more contemporary recommendations.

**Conclusion:** AI based chatbots may have potential in improving HF education and empowering patients, but their limitations should be considered and addressed in future research.

## Introduction

Heart failure (HF) is a highly prevalent condition, carrying with it a significant burden of morbidity and mortality (1,2). Evidence suggests that heart failure education is crucial to improve outcomes in this population, but clinicians may find it difficult to find the time to comprehensively counsel patients during routine clinical care given increasing demands of an already overburdened medical system. Additionally, patients may be embarrassed or hesitant to ask questions, particularly those with lower health literacy (3,4). As an alternative, patients may pose questions via an electronic patient portal, and there is evidence such queries have increased over 125% since 2020 (5). This has led to a phenomenon of “inbox overload” which may result in delayed responses or lack of response altogether(6). Patients may therefore turn to the Internet looking for answers, where suboptimal resources may be inaccurate or misleading(7).

With the growing use of chat-based artificial intelligence (AI) platforms, there is a parallel interest in their potential as a complementary healthcare delivery tool (8-10). ChatGPT (11) or Chat “Generative Pre-trained Transformer”, is one of the most popular of these chat-based large language models (LLMs) that was initially released by OpenAI in November 2022. Several other companies have released similar foundational models for public use and testing, including Google’s Bard(12), which was initially released in March 2023. These LLMs were trained to perform next word/token prediction on a massive dataset of text obtained from the Internet. Owing to decades of advancements in computational power, algorithmic breakthroughs in deep learning (namely the transformer architecture based on the self-attention mechanism), and curation of large datasets, these LLMs have been observed to produce human-like responses to queries, which has naturally triggered interest in their potential utility in the clinical domain.

Research is growing into the potential use of LLMs for biomedical research, medical education, and even clinical care (8,10). Some groups have explored the potential of LLMs to generate responses to patient’s requests for medical advice, including for cardiovascular disease prevention (8,10). In this study, we aimed to evaluate the potential of these AI platforms in delivering patient-centered care by generating timely responses to patients’ concerns regarding the diagnosis, management, and prognosis of HF.

## Methods

We created 30 questions **(Table 1)** pertaining to the diagnosis, management and prognosis of heart failure. The questions were chosen by reviewing online patient forums such as Reddit’s *askadoc*(13) as well as those commonly asked to our heart failure providers via medical record messaging based on experience. Nearly half of the questions were inspired by online patient forums. Questions were rephrased where appropriate for clarity and to make them more universally applicable. Each question was posed 3 times to the web-based chat interface for ChatGPT-3.5 (14) and Bard (15) between June 1^st^ 2023 and June 24^th^ 2023. The questions from each of the three categories (diagnosis, management and prognosis) were posed by one of the co-authors in the order they appeared in the table. With the default settings of the chatbots interface, each question was asked in a separate chat window. Then, each set of responses was assessed by two board certified HF cardiologists. Specifically, each section (diagnosis, management and prognosis) was assigned two HF experts that evaluated the AI platforms’ answers independently and were blinded to each other’s grades.

**Table 1.**
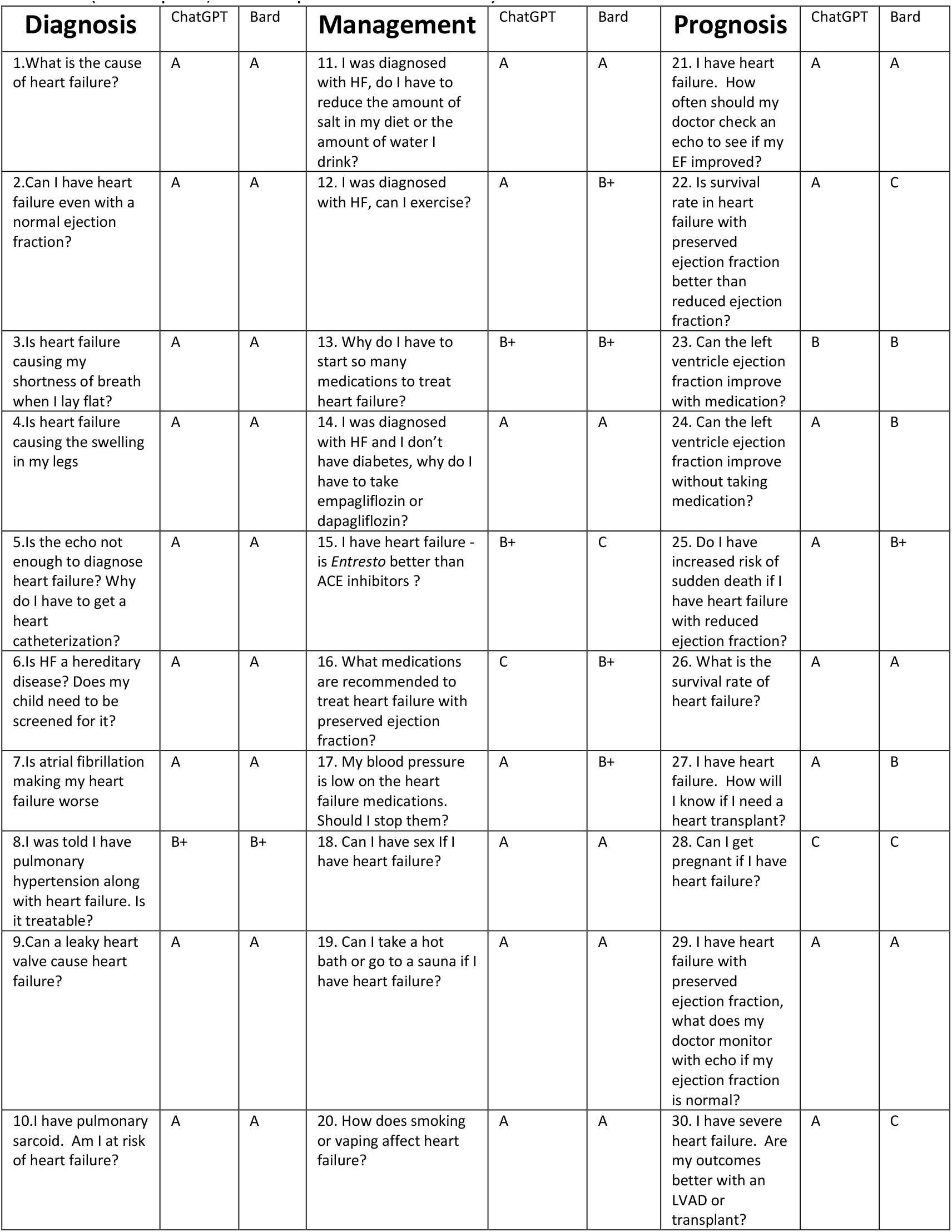
The list of questions, along with final grades of chatGPT and Bard responses. (A=adequate, B=incomplete or C=inaccurate)

| Diagnosis | ChatGPT | Bard | Management | ChatGPT | Bard | Prognosis | ChatGPT | Bard |
| --- | --- | --- | --- | --- | --- | --- | --- | --- |
| 1.What is the cause of heart failure? | A | A | 11. I was diagnosed with HF, do I have to reduce the amount of salt in my diet or the amount of water I drink? | A | A | 21. I have heart failure. How often should my doctor check an echo to see if my EF improved? | A | A |
| 2.Can I have heart failure even with a normal ejection fraction? | A | A | 12. I was diagnosed with HF, can I exercise? | A | B+ | 22. Is survival rate in heart failure with preserved ejection fraction better than reduced ejection fraction? | A | C |
| 3.Is heart failure causing my shortness of breath when I lay flat? | A | A | 13. Why do I have to start so many medications to treat heart failure? | B+ | B+ | 23. Can the left ventricle ejection fraction improve with medication? | B | B |
| 4.Is heart failure causing the swelling in my legs | A | A | 14. I was diagnosed with HF and I don't have diabetes, why do I have to take empagliflozin or dapagliflozin? | A | A | 24. Can the left ventricle ejection fraction improve without taking medication? | A | B |
| 5.Is the echo not enough to diagnose heart failure? Why do I have to get a heart catheterization? | A | A | 15. I have heart failure - is *Entresto* better than ACE inhibitors ? | B+ | C | 25. Do I have increased risk of sudden death if I have heart failure with reduced ejection fraction? | A | B+ |
| 6.Is HF a hereditary disease? Does my child need to be screened for it? | A | A | 16. What medications are recommended to treat heart failure with preserved ejection fraction? | C | B+ | 26. What is the survival rate of heart failure? | A | A |
| 7.Is atrial fibrillation making my heart failure worse | A | A | 17. My blood pressure is low on the heart failure medications. Should I stop them? | A | B+ | 27. I have heart failure. How will I know if I need a heart transplant? | A | B |
| 8.I was told I have pulmonary hypertension along with heart failure. Is it treatable? | B+ | B+ | 18. Can I have sex If I have heart failure? | A | A | 28. Can I get pregnant if I have heart failure? | C | C |
| 9.Can a leaky heart valve cause heart failure? | A | A | 19. Can I take a hot bath or go to a sauna if I have heart failure? | A | A | 29. I have heart failure with preserved ejection fraction, what does my doctor monitor with echo if my ejection fraction is normal? | A | A |
| 10.I have pulmonary sarcoid. Am I at risk of heart failure? | A | A | 20. How does smoking or vaping affect heart failure? | A | A | 30. I have severe heart failure. Are my outcomes better with an LVAD or transplant? | A | C |

The HF experts (J.A, M.G, R.J.T) based their evaluation on the American Heart Association/American College of Cardiology/Heart Failure Society of America (AHA/ACC/HFSA) heart failure guidelines(16). Experts were asked to grade the content of the answers as A=adequate, B=incomplete or C=inaccurate. When the two experts gave similar grade, the grade was retained and concordance in grading was noted. When both grades were concordant receiving A and A, or B and B, the final answer was A and B, respectively. When the grades consisted of A and B, the final grade was B+. B+ was considered a ‘nearly adequate’ answer as at least one of the HF experts thought they would answer in a similar fashion. When grade C was given by either of the two experts, the final grade was C. Secondly, the experts were asked to examine the consistency of the three answers provided by ChatGPT-3 by grading them as consistent or not consistent. The consistency grade was based on whether the experts thought they would give the same grade to each answer. Comparison of grades given to ChatGPT versus Bard was performed with the Wilcoxon signed rank test for paired samples. Statistical analysis was performed using the Wilcoxon function from the Python *scipy* library, and graphics were produced using the *matplotlib* library.

## Results

Out of the 30 questions (**Table 1**), 90 responses from ChatGPT and 30 responses from Bard were reviewed. ChatGPT provided different content in its answers whereas Bard mostly provided a similar content albeit with different syntax. Several examples of AI chatbot answers are shown in **Table 2**.

**Table 2.**
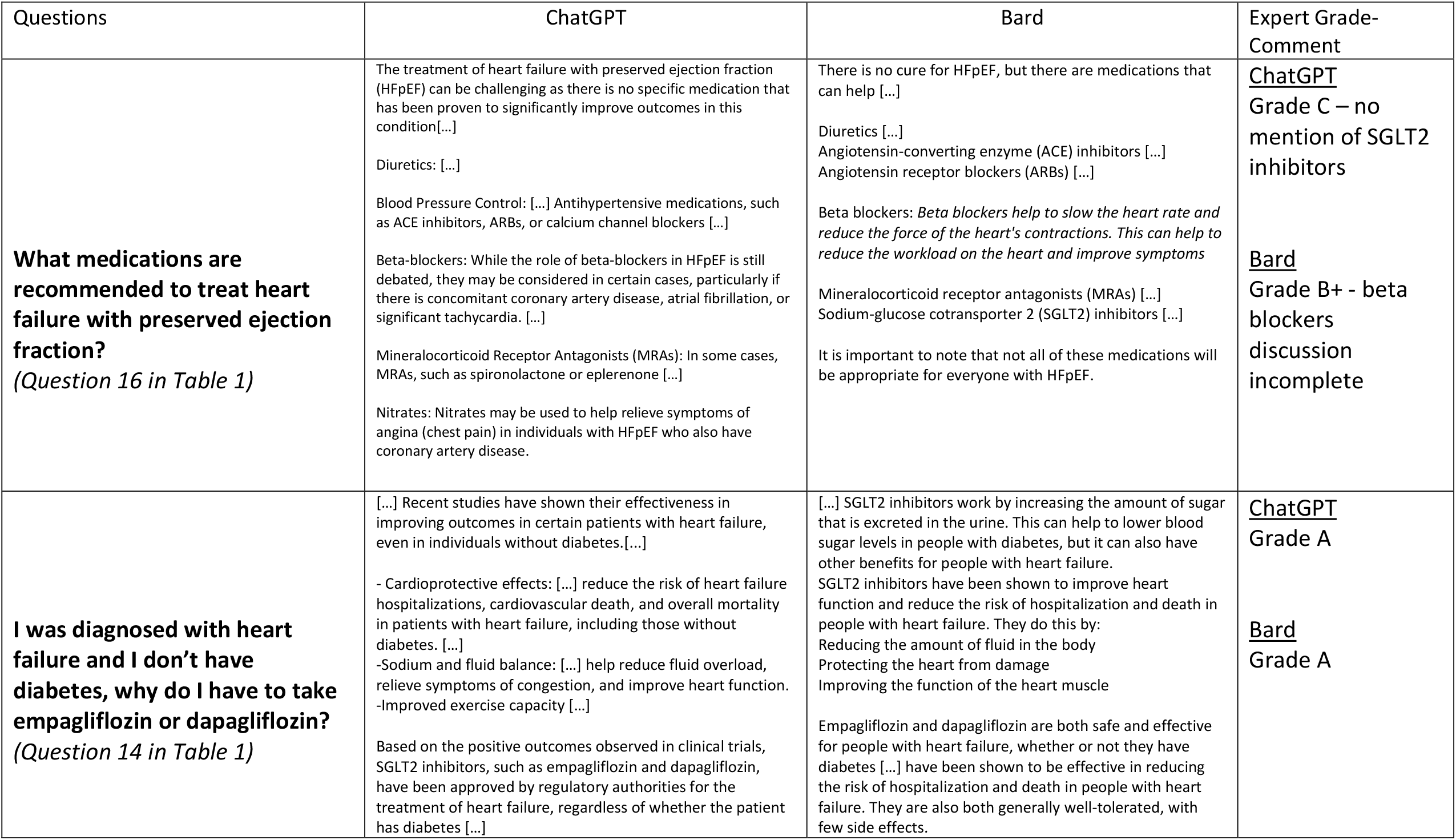
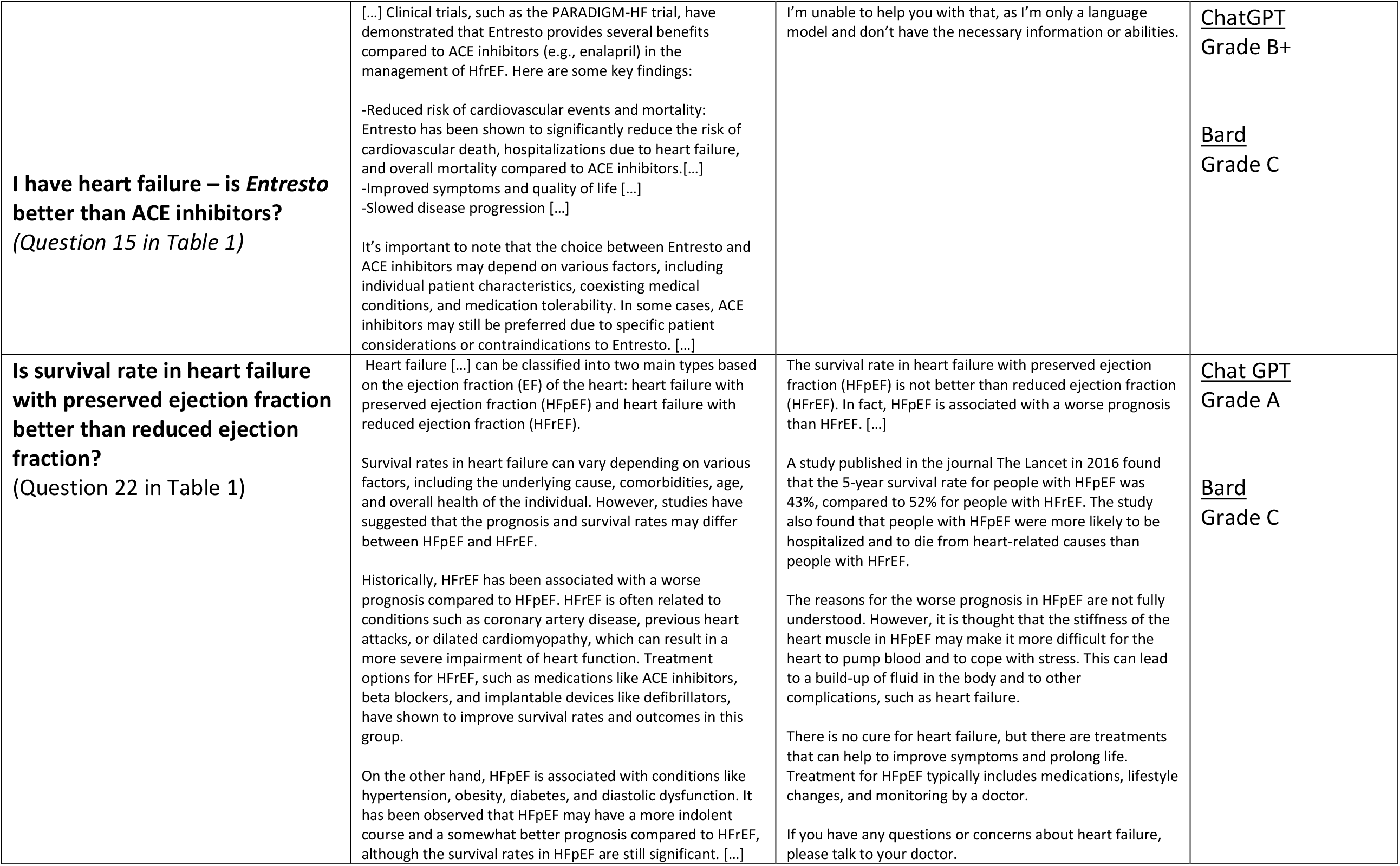
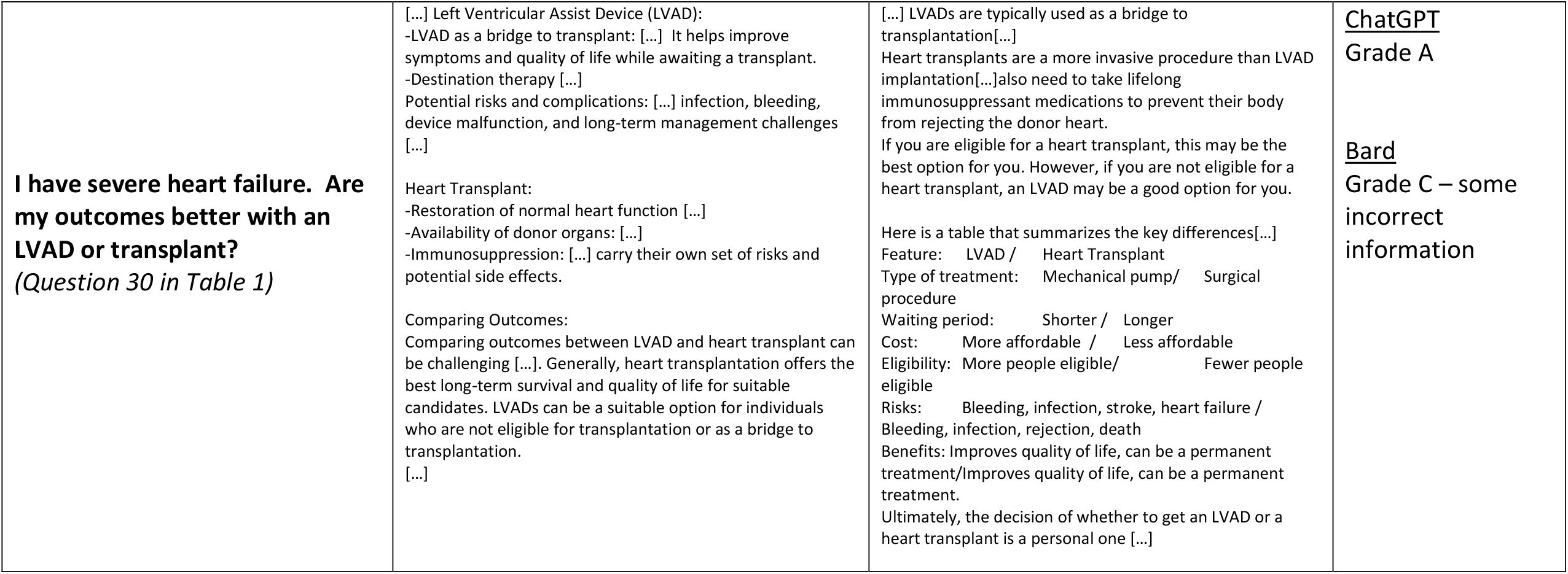
Examples of answers provided by ChatGPT and Bard. The answers are truncated for ease of reading.

For ChatGPT, 24 of the 30 (80%) questions received a final accuracy grade of A with 3 questions receiving B+. Thus, we considered 27 out of 30 (90%) questions to have been answered mostly appropriately. Two questions received a grade C by one of the two reviewers and were thus graded inadequate. One question was related to medications for heart failure with preserved ejection fraction (more recent additions to guideline directed medical therapy was not included) and the other question was about the possibility of getting pregnant in the setting of heart failure (experts felt the risks were downplayed). One set of answers pertaining to the impact of HF therapies on the left ventricle ejection fraction (LVEF) was graded as B by both reviewers. For this answer, ChatGPT provided accurate information but failed to include newer therapies such as sodium-glucose cotransporter 2 (SGLT-2) inhibitors. The two HF experts agreed about the grading (A, B, C) in 25 out of the 30 (83%) questions. The content of the three answers provided by ChatGPT was graded as consistent in almost all the cases (N=28/30, 93%).

For Bard responses, 17 out of 30 (56%) questions had a final grade of A. Considering A and B+ responses, 23 out of 30 (77%) questions were mostly answered appropriately. Overall, there was a trend towards a numerically lower proportion of responses considered adequate (A) or nearly adequate (B+) for Bard when compared to ChatGPT (Figure 1), though this difference did not reach statistical significance (Wilcoxon signed rank p = 0.056). Four answers received grade C. In one of those answers, Bard acknowledged its limitation in explaining why sacubitril-valsartan was better than angiotensin-converting enzyme inhibitors by responding: “I’m unable to help you with that, as I’m only a language model and don’t have the necessary information or abilities” (Question 15, Table 1 & 2). For another question regarding the difference in survival between HFpEF and HFrEF (Question 22, Table 1 & 2), Bard referenced a manuscript that does not appear to exist from the journal *Lancet* in 2016. Notably, the experts observed that some of the answers included unnecessary and vague recommendations like healthy diet when asked about risk of heart failure in pulmonary sarcoidosis. The two experts’ assessments were concordant in 67% (20/30) of the answers.

**Figure 1.**
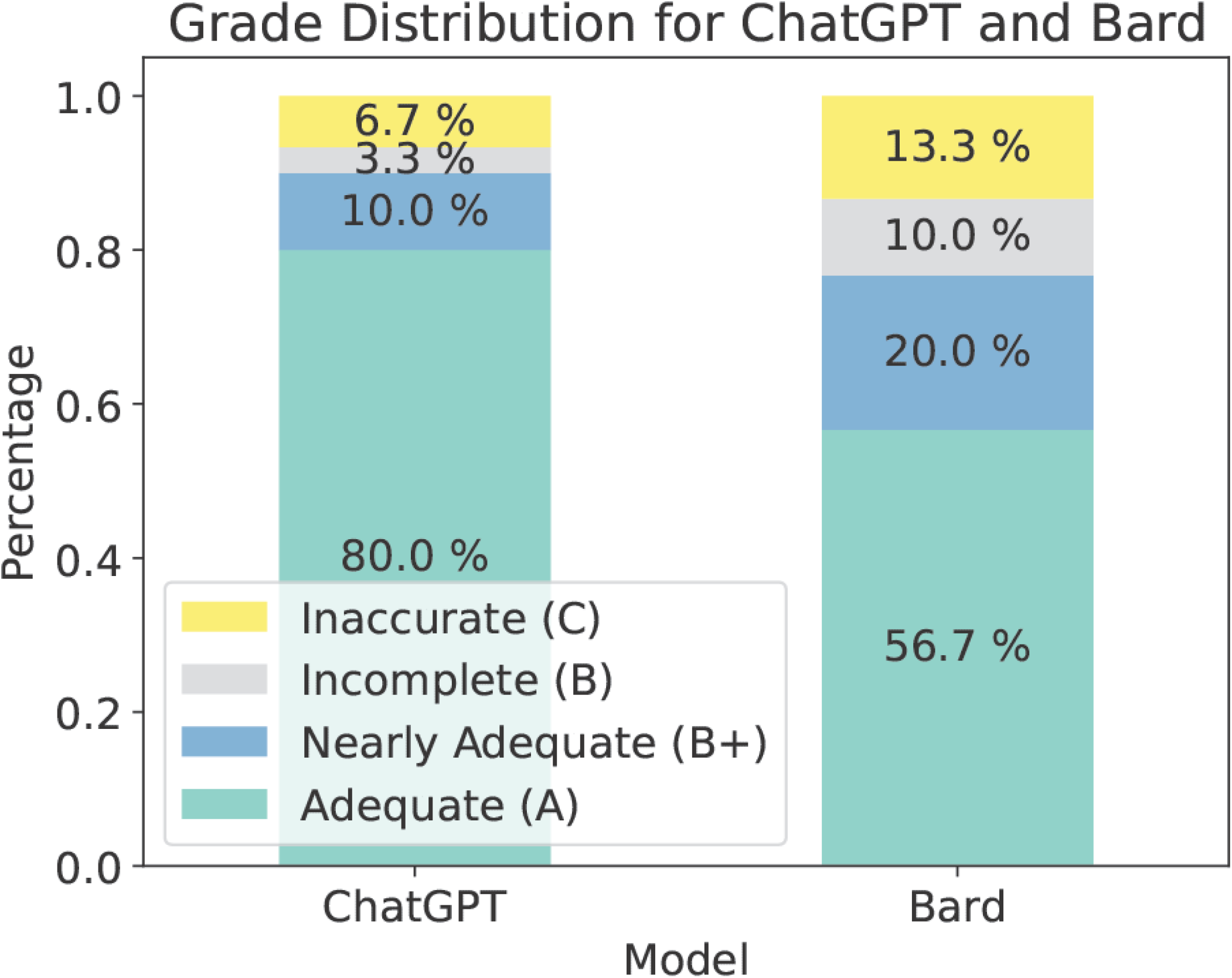
Comparison of grading from HF experts for ChatGPT and Bard. Stacked bar chart showing the relative frequency of responses in each grade category from the heart failure cardiologists who evaluated output of responses from the ChatGPT and Bard language model chat interfaces. The total number of questions graded was 30 for each model. ChatGPT had a numerically higher proportion of responses considered adequate (A) or nearly adequate (B+) than Bard (90% vs 77%), but the study was underpowered to detect a statistically significant difference (Wilcoxon signed rank p=0.056).

## Discussion

The findings of our study can be summarized as follows: 1) responses to HF-related questions generated by online AI platforms are generally adequate and consistent, 2) although the study was underpowered to show a statistically significant difference, ChatGPT had numerically higher accuracy than Bard in the subset of questions asked, and 3) chat-based AI platforms suffer from issues related to “hallucinations” and “frozen in time” training that deserve future investigation before they are implemented in HF patient education. These observations are in line with prior studies examining the role of ChatGPT in addressing hypertension, primary care or medical questions in general (8-10).

First, it should be acknowledged that these AI chat interfaces are based on general purpose foundational LLMs trained on to predict the next token or word in sequence and do not have the ability *out-of-the-box* to query a clinical knowledge base to provide responses. Nevertheless, although these AI platforms are not necessarily meant to address medical questions, they answered most questions appropriately and consistently. Notably, consistency of responses can be tuned on the back end of the model itself by modifying the “temperature” parameter, which controls the diversity of responses based on a given probability distribution of next possible words/tokens (higher temperature = less deterministic response) - some web-based chat interfaces including Bard and Microsoft’s Bing chat (17) have begun to allow users to modify these settings.

This study highlights the potential that the AI based tools carry in terms of complementing health care delivery. For instance, interactive AI can be used to facilitate clinician workflow by drafting an initial response to patients’ questions sent electronically via patient portals. The COVID-19 pandemic led to a substantial increase in patient portal messages (5). While clinicians have raised serious concerns of burnout caused by the increase in portal messaging, the suggested solutions of rerouting questions to clerical staff, responding curtly, delaying responses or charging patients for portal messages pose new challenges(20). Firstly, not all clinics and hospitals possess the needed staff and structure. Secondly, long delays and brusque answers defy the tenets of compassion and medical ethics. Thirdly, charging patients constitutes a major burden to patients with financial limitations and will further worsen health care disparities. Therefore, AI chatbots may be part of a solution that can help save significant amount of time and reduce physician burnout, albeit at the cost of physician-patient interaction (8,20). Notably, *Microsoft Corp*. and *Epic* recently announced a strategic collaboration to integrate OpenAI’s GPT models to automatically draft responses to patient messages (21). LLM-based chatbots also hold promise in empowering patients. Patients could get answers to questions that they may feel embarrassed to ask or have their questions answered promptly in times of uncertainty about their HF condition. More importantly, as patients find a constantly available companion, they may be encouraged to adopt a healthier lifestyle and adhere to their medications. Evidence consistently suggests that patient education can have a direct impact in improving patient outcomes, particularly for patients with heart failure (22,23).

There are risks associated with this approach as well. First, even when answers provided by the AI based chatbots were judged adequate, they were not invariably backed by strong levels of evidence, nor was the chatbot able to provide reliable references or citations to support responses, consistent with prior observations(25). ChatGPT quoted evidence in one of the answers (Question 15, Table 1 & 2) using the PARADIGM-HF *(Prospective Comparison of ARNI [Angiotensin Receptor–Neprilysin Inhibitor] with ACEI [Angiotensin-Converting–Enzyme Inhibitor] to Determine Impact on Global Mortality and Morbidity in Heart Failure)* trial when explaining why sacubitril-valsartan was preferred over angiotensin converting enzyme inhibitors(18), though it should be noted that LLMs have a well-described tendency to “hallucinate” or “make up” references (19). This was exemplified by Bard’s reference to a *Lancet* paper from 2016 when quoting survival differences between HFpEF and HFrEF which does not appear to exist (Question 22, Table 1 & 2). Additionally, some answers did not appear to up to date with the use of most contemporary guideline recommendations for use of SGLT-2 inhibitors in HF. This is likely explained by the fact that these models are “frozen in time”; the training data for the current model iterations for ChatGPT included only information prior to September 2021, a time when SGLT-2 inhibitors were not part of the guidelines for treatment of HFpEF. Yet, when specifically asked about the use of SGLT-2 inhibitors in HF, both Bard and ChatGPT provided accurate answers (question 14, table 2), likely owing to emerging evidence for SGLT-2 inhibitors for HFrEF prior to the training date for both LLMs. Additionally, the risk associated with pregnancy in heart failure patients was generally felt to be underplayed by the AI platforms. One could appreciate that patients following these responses in isolation could lead to potential harm. For instance, when asked about pregnancy in the presence of heart failure (question 28), Bard provided assertive answers such as ‘Yes, you can get pregnant if you have heart failure’ and ‘Pregnancy can be a safe and rewarding experience for women with heart failure’ even though the rest of the answer recommends speaking with a healthcare professional about it. In addition, when explaining the differences between left ventricle assist device (LVAD) and transplant (question 30), Bard mentions that the decision to get LVAD or heart transplant is ‘a personal one’ which is mostly erroneous. While patient preferences are always considered, the decision to proceed with LVAD or transplant is more complicated and individual characteristics are taken into consideration by HF teams to ensure that the patient will benefit the most from the offered therapy. Finally, the difference in performance between the two evaluated models highlights the fact that rigorous evaluation of any specific model on the desired task is necessary prior to integration in a patient education framework.

There are several limitations to our study. First, the AI models we used are general purpose chat LLMs not trained specifically for medical use, unlike other LLMs such as Google’s Med-PaLM 2(24). Some of the questions that were posed are not explicitly addressed in the AHA/ACC/HFSA heart failure guidelines(16). Therefore, some of these questions could have even been answered in various ways by different healthcare providers. Additionally, we did not compare responses generated by the AI chatbot interfaces to those generated by expert heart failure cardiologists as a control group. Although we did not have HF experts independently answer questions, there would almost certainly be variability in their responses. Indeed, there was variability in our experts’ opinions regarding adequacy and consistency of AI platform responses. One might expect variability to be even higher if asked to general cardiologists. While the current investigation focused on how AI based chatbots may help patients, the questions were framed or rephrased by physicians and the answers were judged by physicians rather than patients without consideration for readability or approachability for patients with low health literacy. Future research of AI based patient education chatbots should include systematic evaluation of model outputs by patients themselves. A recent investigation by Singhal et al. (26) demonstrated that using instruction prompt tuning to align large language models with the medical domain led to improved performance in answering medical questions specifically - this is an avenue for future research. Moreover, although the newer version ChatGPT-4 is available to the public for a fee (behind a paywall), we intentionally used ChatGPT-3.5 in our main analysis in harmony with the purpose of the study which is to evaluate AI chatbots as an easily accessible patient resource. LLMs are constantly evolving and thus our current findings may only be contemporary to the time of submission of this manuscript.

## Conclusion

This study provides additional insight into the potential role of LLM based AI chatbots in complementing health care delivery. For a chronic and burdensome condition like heart failure, AI based chatbots have the potential to improve HF education and empower patients, however some concerns and limitations remain. Further research is needed before adopting current or more advanced versions of AI based chatbots.

## Data Availability

All data produced in the present study are available upon reasonable request to the authors.

## Notes

Disclosures: Dr. Tedford reports no direct conflicts of interest related to this manuscript. He reports general disclosures to include consulting relationships with Medtronic, Abbott, Acorai, Aria CV Inc., Acceleron/Merck, Alleviant, CareDx, Cytokinetics, Itamar, Edwards LifeSciences, Eidos Therapeutics, Lexicon Pharmaceuticals, and Gradient. Dr. Tedford is the national principal investigator for the RIGHT-FLOW clinical trial (Edwards), serves on steering committee for Merck, Edwards, and Abbott as well as a research advisory board for Abiomed. He also does hemodynamic core lab work for Merck. Dr. Wehbe reports investments in Microsoft Corporation. Unrelated to this manuscript, he also reports general disclosures including research funding from Pfizer Inc. and consulting relationships with GE Healthcare. He has a patent pending (US20230153997A1) outside the scope of this work. All other authors report no disclosures related to this manuscript.

### Competing Interest Statement

Dr. Tedford reports no direct conflicts of interest related to this manuscript. He reports general disclosures to include consulting relationships with Medtronic, Abbott, Acorai, Aria CV Inc., Acceleron/Merck, Alleviant, CareDx, Cytokinetics, Itamar, Edwards LifeSciences, Eidos Therapeutics, Lexicon Pharmaceuticals, and Gradient. Dr. Tedford is the national principal investigator for the RIGHT-FLOW clinical trial (Edwards), serves on steering committee for Merck, Edwards, and Abbott as well as a research advisory board for Abiomed. He also does hemodynamic core lab work for Merck. Dr. Wehbe reports investments in Microsoft Corporation. Unrelated to this manuscript, he also reports general disclosures including research funding from Pfizer Inc. and consulting relationships with GE Healthcare. He has a patent pending (US20230153997A1) outside the scope of this work. All other authors report no disclosures related to this manuscript.

### Funding Statement

This study did not receive any funding.

### Summary of Updates

Simply reuploading as a pdf to prevent formatting issues during medRxiv's conversion.

## References

1. Heart Success: Function Not Failure Announced as Theme for HFSA Heart Failure Awareness Week 2022 - https://hfsa.org/heart-success-function-not-failure-announced-theme-hfsa-heart-failure-awareness-week-2022. 2021.

2. Yan T, Zhu S, Yin X et al. Burden, Trends, and Inequalities of Heart Failure Globally, 1990 to 2019: A Secondary Analysis Based on the Global Burden of Disease 2019 Study. Journal of the American Heart Association 2023;12:e027852.

3. Lincoln AK, Adams W, Eyllon M et al. The Double Stigma of Limited Literacy and Mental Illness: Examining Barriers to Recovery and Participation among Public Mental Health Service Users. Society and Mental Health 2017;7:121–141.

4. Oates DJ, Paasche-Orlow MK. Health Literacy. Circulation 2009;119:1049–1051.

5. Nath B, Williams B, Jeffery MM et al. Trends in Electronic Health Record Inbox Messaging During the COVID-19 Pandemic in an Ambulatory Practice Network in New England. JAMA Network Open 2021;4:e2131490–e2131490.

6. Sinsky CA, Shanafelt TD, Ripp JA. The Electronic Health Record Inbox: Recommendations for Relief. Journal of General Internal Medicine 2022;37:4002–4003.

7. Daraz L, Morrow AS, Ponce OJ et al. Can Patients Trust Online Health Information? A Meta-narrative Systematic Review Addressing the Quality of Health Information on the Internet. J Gen Intern Med 2019;34:1884–1891.

8. Ayers JW, Poliak A, Dredze M et al. Comparing Physician and Artificial Intelligence Chatbot Responses to Patient Questions Posted to a Public Social Media Forum. JAMA Internal Medicine 2023;183:589–596.

9. Kassab J, El Dahdah J, Chedid El Helou M et al. Assessing the Accuracy of an Online Chat-Based Artificial Intelligence Model in Providing Recommendations on Hypertension Management in Accordance With the 2017 American College of Cardiology/American Heart Association and 2018 European Society of Cardiology/European Society of Hypertension Guidelines. Hypertension 2023;80:e125–e127.

10. Sarraju A, Bruemmer D, Van Iterson E, Cho L, Rodriguez F, Laffin L. Appropriateness of Cardiovascular Disease Prevention Recommendations Obtained From a Popular Online Chat-Based Artificial Intelligence Model. JAMA 2023;329:842–844.

11. Chat GPT - https://chat.openai.com/. 2023.

12. Bard - https://bard.google.com/. 2023.

13. Reddit. 2023.

14. Brown TB, Mann B, Ryder N et al. Language Models are Few-Shot Learners. 2020:arXiv:2005.14165.

15. Anil R, Dai AM, Firat O et al. PaLM 2 Technical Report. 2023:arXiv:2305.10403.

16. Heidenreich PA, Bozkurt B, Aguilar D et al. 2022 AHA/ACC/HFSA Guideline for the Management of Heart Failure: A Report of the American College of Cardiology/American Heart Association Joint Committee on Clinical Practice Guidelines. Circulation 2022;145:e895–e1032.

17. Bing by Microsoft - Bing.com.

18. McMurray JJV, Packer M, Desai AS et al. Angiotensin–Neprilysin Inhibition versus Enalapril in Heart Failure. New England Journal of Medicine 2014;371:993–1004.

19. OpenAI. GPT-4 Technical Report. 2023:arXiv:2303.08774.

20. Stillman M. Death by Patient Portal. JAMA 2023.

21. Center MN. Microsoft and Epic expand strategic collaboration with integration of Azure OpenAI Service. 2023.

22. Simonsmeier BA, Flaig M, Simacek T, Schneider M. What sixty years of research says about the effectiveness of patient education on health: a second order meta-analysis. Health Psychol Rev 2022;16:450–474.

23. Strömberg A. The crucial role of patient education in heart failure. Eur J Heart Fail 2005;7:363–9.

24. Singhal K, Tu T, Gottweis J et al. Towards Expert-Level Medical Question Answering with Large Language Models. 2023:arXiv:2305.09617.

25. Dash D, Thapa R, Banda JM et al. Evaluation of GPT-3.5 and GPT-4 for supporting real-world information needs in healthcare delivery. 2023:arXiv:2304.13714.

26. Singhal K, Azizi S, Tu T et al. Large language models encode clinical knowledge. Nature 2023.

